# LLM Instruction-Example Adaptive Prompting (LEAP) Framework for Clinical Relation Extraction

**DOI:** 10.1101/2023.12.15.23300059

**Authors:** Huixue Zhou, Mingchen Li, Yongkang Xiao, Han Yang, Rui Zhang

**Author notes:** Dr. Rui Zhang, PhD Division of Computational Health Sciences, Department of Surgery University of Minnesota 11-132 Phillips-Wangensteen Building, 516 Delaware St SE, Minneapolis, MN 5545.

## Abstract

**Objective:** To investigate the demonstration in Large Language Models (LLMs) for clinical relation extraction. We focus on examining two types of adaptive demonstration: instruction adaptive prompting, and example adaptive prompting to understand their impacts and effectiveness.

**Materials and Methods:** The study unfolds in two stages. Initially, we explored a range of demonstration components vital to LLMs’ clinical data extraction, such as task descriptions and examples, and tested their combinations. Subsequently, we introduced the Instruction-Example Adaptive Prompting (LEAP) Framework, a system that integrates two types of adaptive prompts: one preceding instruction and another before examples. This framework is designed to systematically explore both adaptive task description and adaptive examples within the demonstration. We evaluated LEAP framework’s performance on the DDI and BC5CDR chemical interaction datasets, applying it across LLMs such as Llama2-7b, Llama2-13b, and MedLLaMA_13B.

**Results:** The study revealed that *Instruction + Options + Examples* and its expanded form substantially raised F1-scores over the standard *Instruction + Options* mode. LEAP framework excelled, especially with example adaptive prompting that outdid traditional instruction tuning across models. Notably, the MedLLAMA-13b model scored an impressive 95.13 F1 on the BC5CDR dataset with this method. Significant improvements were also seen in the DDI 2013 dataset, confirming the method’s robustness in sophisticated data extraction.

**Conclusion:** The LEAP framework presents a promising avenue for refining LLM training strategies, steering away from extensive finetuning towards more contextually rich and dynamic prompting methodologies.

## INTRODUCTION

Clinical relation extraction (RE), a natural language processing task, has emerged as a crucial task within healthcare informatics due to its significant role in deciphering drug interactions, side effects, and treatment outcomes [1]. The evolution of this field has been largely influenced by the advent of transformer-based models such as BERT, along with its specialized variants like BioBERT, BiolinkBERT, SciBERT, and PubmedBERT, which have established new performance benchmarks [2–6].

The rise of Generative Large Language Models (LLMs) has generated considerable interest in their application within clinical contexts, primarily due to their versatility and adaptability in processing complex medical data [7–14]. These models distinguish themselves from previous pre-trained models, such as BERT, by incorporating instructions into their input, providing explicit guidance for task completion, and generating expected responses. Typically, the input for generative LLMs consists of a task description followed by input data, often supplemented with examples to serve as demonstration.

Current research on generative LLMs places a significant focus on manual instruction crafting or instruction tuning, aimed at enhancing the efficiency of LLMs across diverse NLP tasks [15–18]. More sophisticated techniques have emerged, such as the automated search methods developed by Prasad et al. [19] and Wei et al. [20], which identify optimal instructions or word choices for generative LLM instruction.

Technological advancements like Chain-of-Thought (CoT) and in-context learning have significantly propelled the development of generative LLMs. The CoT methodology has particularly been a game-changer, encouraging LLMs to unfold complex reasoning step by step, which enhances their ability to handle multifaceted tasks. Several Studies [21–14] have highlighted CoT’s effectiveness in promoting such detailed reasoning processes within generative LLMs. In a similar vein, in-context learning has also proven to be instrumental. By strategically providing LLMs with pertinent information, such as related entities [22,25,26] and illustrative examples [26,27], this approach improves models’ contextual understanding and their application of knowledge to RE.

Additionally, emergence of retrieval based LLMs represents a cutting-edge and promising development in the field of generative LLMs for RE. This strategy gains its strength from synergizing with in-context learning, aiming to access and incorporate highly relevant supplementary resources, thereby enriching the model’s comprehension and analytical capabilities [27–29]. These studies illustrate the transformative impact of combining retrieval methodologies with generative LLMs, effectively enhancing their performance by providing them with a more nuanced and contextually rich knowledge base.

Despite these advancements, challenges persist in crafting manual instruction or retrieval-based demonstrations. Developing high-quality instructions that capture intended behaviors is complex, and instruction datasets often lack size, diversity, and creativity [30]. While in previous study, when the manually crafted prompt meet its limit in transformer-based models, studies introduced a more dynamic method, soft prompt, which involves learning a prompt embedding matrix to dynamically adapt the PLMs [31–33].

Building upon the concept of the soft prompt, our work introduces an Instruction-Example Adaptive Prompting Framework. This framework dynamically learns the embedding of instructions and examples to adapt the generative LLMs for clinical relation extraction. This study is driven by the goal of optimizing the use and application of demonstrations within LLMs to fully exploit their capabilities in this complex domain. Our study is anchored on two primary contributions:

1. Demonstration Diversity Operation: We designed various demonstration components and undertook a comprehensive analysis to ascertain their impact on enhancing LLM performance in clinical relation extraction tasks. This part of our research involves defining and experimenting with different configurations of demonstration elements to determine the most effective combinations of demonstration elements for relation extraction task.
2. Development of LLM Instruction-Example Adaptive Prompting (LEAP) Framework: This framework is specifically designed to assess the impact of demonstration clarity on LLM performance. By focusing on the nuances of task descriptions and the integration of examples, the LEAP framework aims to provide deeper insights into how instructional design and in-context examples influences the accuracy and efficiency of LLMs in parsing and understanding complex clinical relations.

To the best of our knowledge, LEAP framework is the first framework to implement adaptive prompt in demonstration of generative LLMs. Through these contributions, our study seeks to push the boundaries of current understanding and application of LLMs in clinical relation extraction, particularly in deciphering intricate chemical-chemical interactions.

## BACKGROUND

### Soft prompt

In the realm of pre-trained language models (PLMs), fine-tuning (FT) has historically been the go-to method, which involves the updating of all model parameters for a specific task. However, as the scale of models expands rapidly, alternative training strategies like prompting and prompt-oriented fine-tuning have gained prominence [34]. Prompting entails freezing all parameters of a PLM and utilizing a natural language prompt to query the model [35]. In the concept of soft prompt tuning, only continuous prompts are adjusted. Pioneered by Liu et al. [31], Lester et al. [32], and by Liu et al. [33], this involves adding trainable continuous embeddings, also known as continuous prompts, to the input word embeddings sequence.

### Instruction tuning (InsT)

A major challenge with generative LLMs is the mismatch between their training goals and user requirements [30]. To address this issue, InsT has become a crucial approach. InsT improves the capabilities and control of LLMs by training them with (DEMONSTRATION+INPUT SENTENCE, OUTPUT) pairs. The DEMONSTRATION here includes instruction and examples: Instruction refers to the user’s directive, these instructions act as a guiding framework, aligning the model’s outputs with the desired response criteria or specific domain knowledge, thereby giving users the ability to direct the model’s responses. While OUTPUT is the model’s ideal response adhering to this instruction. For instance, in a RE task, an Instruction might state, “You are an excellent linguist. The task is to predict relationship between the given head entity and tail entity within a given sentence, this relation which must be in (‘mechanism’, ‘effect’, ‘advice’, ‘int’, ‘None’).” The examples within a demonstration typically include an input sentence paired with its desired output, illustrating the model’s target response. For example, “Input: In the sentence: Milk, milk products, and calcium-rich foods or drugs may impair the absorption of EMCYT. The relationship between calcium and EMCYT is? Response: mechanism.”

## METHODS

### Overview of methods

Our methodology is structured into two phases. In the initial phase, we concentrated on investigating various combinations of demonstration components, which is a fundamental step toward the phase II. This exploration aimed to identify the most effective arrangements of these demonstration components in guiding LLMs for clinical relation extraction tasks. The second phase involved the development and implementation of the LEAP framework. This framework was designed to facilitate a more dynamic interaction between the LLM and the demonstration, allowing for an adaptive approach to both the instructions and the examples provided to the LLM models. Through this method, we aimed to comprehensively assess and enhance the efficiency and accuracy of LLMs in clinical relation extraction. In the following sections, we will introduce the dataset and task, phase I: Demonstration Diversity Operation, and phase II: Implement of LEAP framework, and Experiments and evaluation.

### Datasets and task formulation

DDI Extraction 2013 Corpus (DDI 2013): This dataset, introduced by Herrero-Zazo [36], is used for the drug-drug interaction task. The training dataset of DDI extraction consists of 714 texts (572 from DrugBank and 142 MedLIne abstracts) and test dataset consists of 158 DrugBank Texts and and 33 MedLine abstracts. We implement a train/validation/test split of 543/181/181 files with those texts, respectively.

BC5CDR: Introduced by Taboureau et al. [37], BC5CDR is a dataset curated for chemical-disease relation extraction tasks. It contains 1,500 documents evenly split into training, validation, and test sets, each consisting of 500 documents.

We deploy LLMs for the purpose of RE, with the goal of identifying the relationship between two designated entities within a given text context. The input for the LLM consists of a context labeled as 𝐶, incorporating a narrative passage p containing two entities, head entity/subject E1 and tail entity/object E2. The LLM’s role is to interpret the context and produce a descriptive relation r that accurately reflects the connection between the entities. The output is expected to be a string or label that categorizes the relationship between E1 and E2, adhering to a pre-defined set of relationship categories. In cases where the entities’ relationship does not correspond to any predefined categories, the LLM should output “NULL” or an equivalent term.

### Demonstration diversity operation

In this section, we delineate the constituent elements of demonstration and the various demonstration modes and examine their impact on the performance of LLMs in clinical RE tasks. The foundational structure of a demonstration here is composed of a task description and options. The task description explicates the nature of the task, setting the stage for the LLM’s operation. The options component enumerates all possible relations as defined by the corresponding dataset, which is represented in Figure 2. Furthermore, we have enriched the demonstration with additional components to enhance the RE process. A key addition is the inclusion of option descriptions, which serve to elucidate clinical entity relationships that may not be immediately apparent to the LLM. For instance, the relation type “int” within the DDI dataset could be perplexing for the LLM, as it refers a confirmed drug-drug interaction that, without further detail, remains ambiguous.

**Figure 1.**
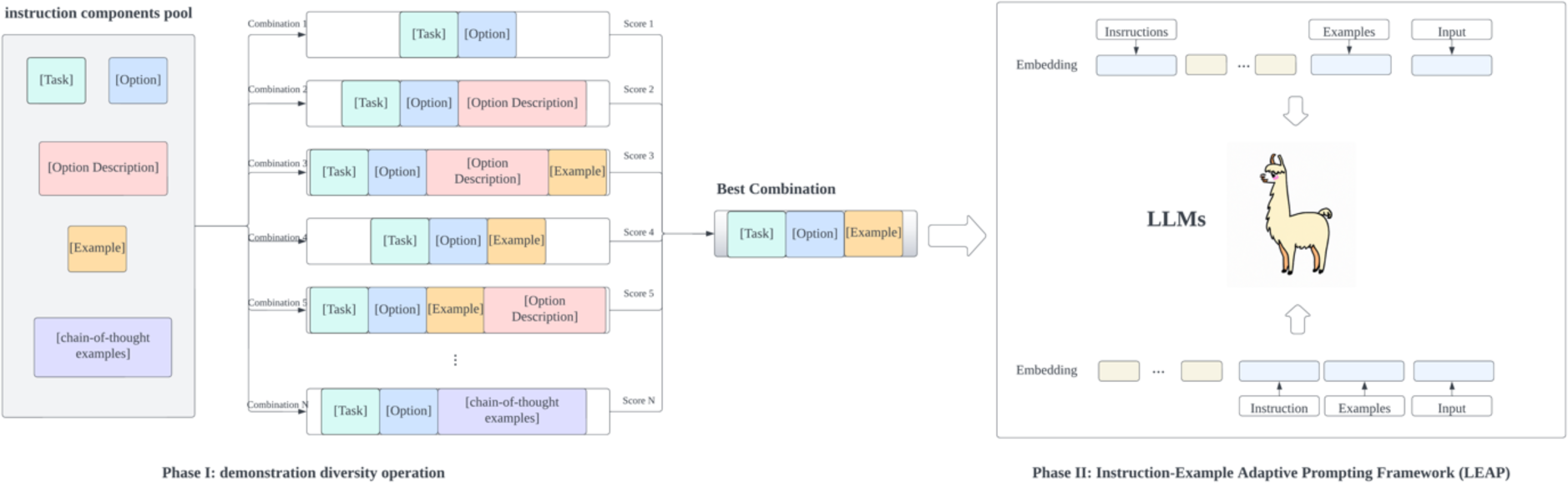
Overview of Study.

**Figure 2.**
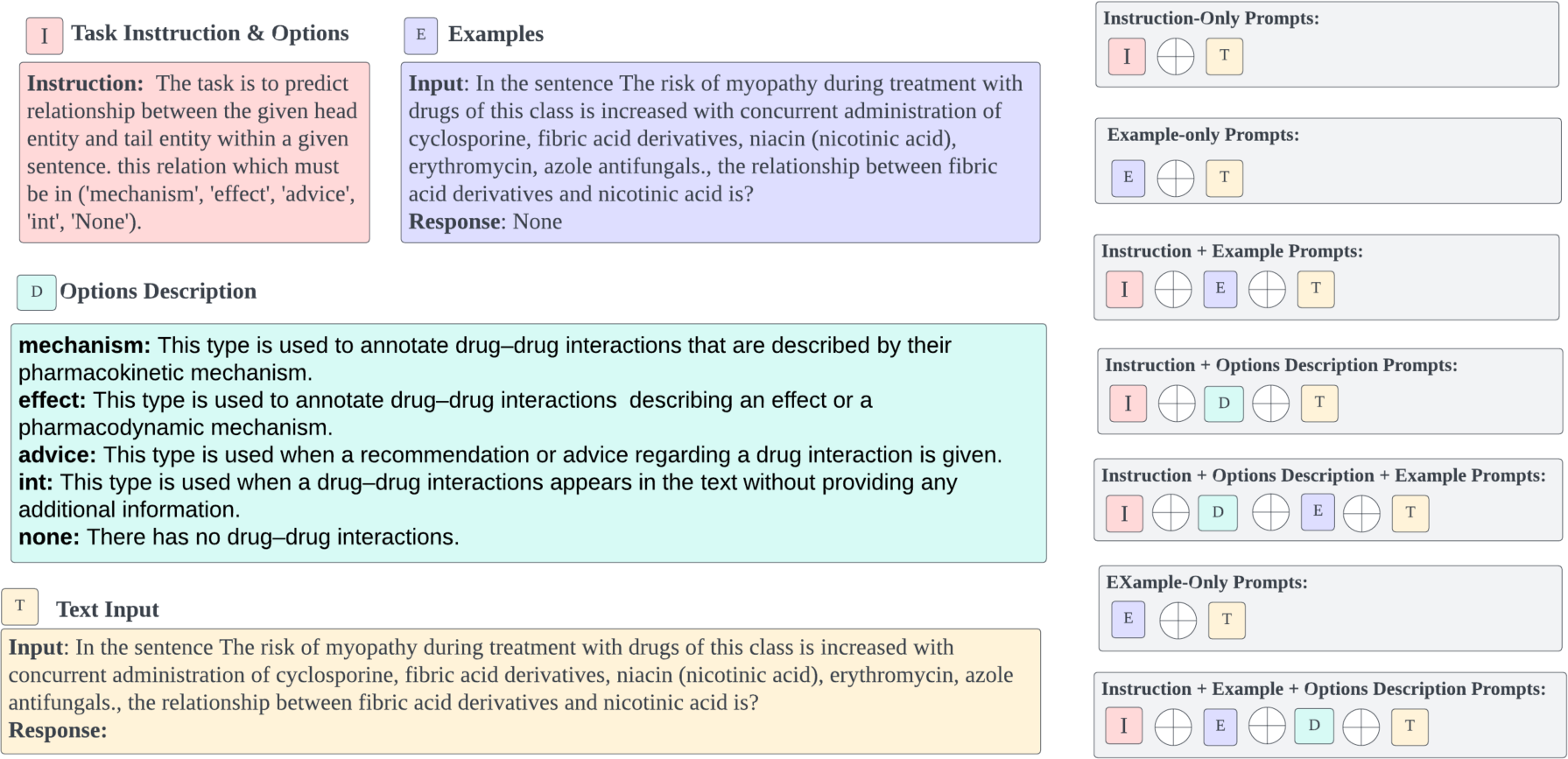
Example of Instruction Searching.

Another crucial component is the inclusion of examples, which supply supplementary in-context information, thereby enhancing the LLM’s comprehension of the task at hand. The demonstration design also accounts for the potential influence of reasoning CoT methodologies, which further require the LLMs to provide extra explanation for the relation of entities. Subsequently, we constructed various prompt modes through diverse amalgamations of these instructional components, each designed to test and optimize the LLM’s performance in accurately extracting clinical relations. This systematic approach allowed us to measure the impact of different instructional designs and set the stage for developing the LLM Instruction-Example Adaptive Prompting Framework (LEAP) in the subsequent phase of the study.

### Implement of Instruction-Example Adaptive Prompting (LEAP) Framework

In the traditional instruction tuning shown in Figure 3a, the *Instruction + Example* demonstration and input sentence are fed into the LLM to generate the relation type between two entities in a sentence. The instructions and examples are consistently crafted in natural language. To enable demonstration dynamically to update the LLM and mitigate input limitations arising from lengthy examples, we introduce LEAP framework. LEAP is designed to enhance the generative LLM’s output by strategically integrating adaptive prompts into the input context. This approach assists the model in generating more precise relational outputs.

**Figure 3.**
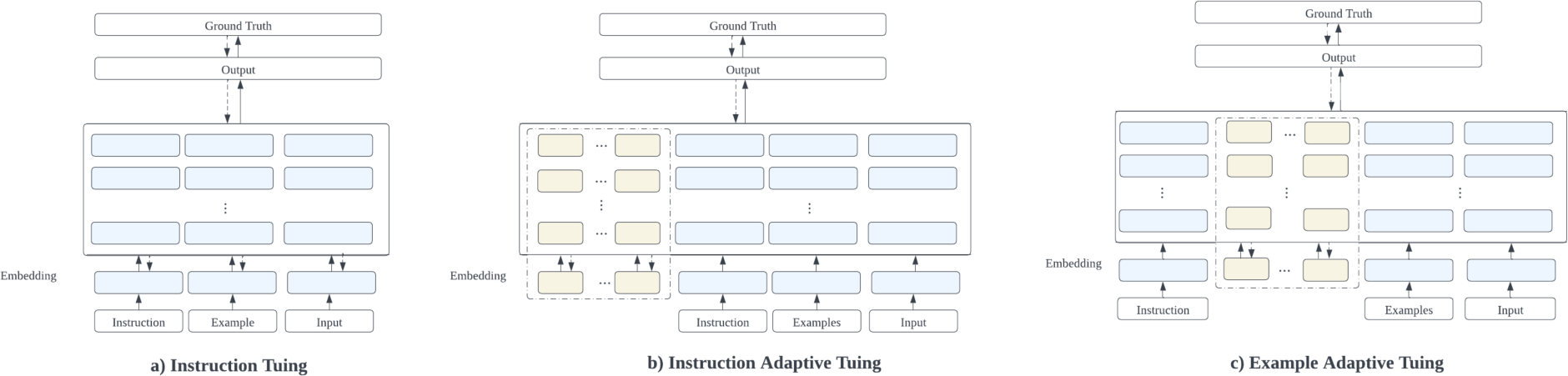
Instruction tuning (a) and LLM Instruction-Example Adaptive Prompting (b, c). Figure 4. performance of demonstration diversity operation.

Given the embedding of *Instruction + Example* demonstration *E* = {*I*_1_, … *I_j_*, *e*_1_, …, *e_n_* } where *I_j_* denotes the token embedding for each token in the instruction, and *e_n_* denotes the token embedding for each token in the randomly sampled example from a valid dataset. We crafted two distinct methods for integrating these adaptive prompts:

1. Instruction Adaptive Tuning Method (Figure 3b): This method involves the insertion of soft vectors before the task description. The demonstration sequence embedding is represented 𝐸 = {𝑣_1_, …, 𝑣_*i*_, …, 𝐼_1_, …, *I_j_*, …, 𝑒_1_, …, *e_n_*} where 𝑣_*i*_ denotes the soft prompt vectors, which is randomly initialized from the embedding layer 𝑀 of LLM and is learnable during the training process. *I_j_* denotes the token embedding for each token in the instruction. 𝑒_n;_ denotes the token embedding for each token in the randomly sampled example from valid dataset. This configuration is designed to prime the LLM with additional task-relevant information before it processes the actual context.
2. Example Adaptive Tuning Method (Figure 3c): The second method positions the soft vectors between the task description and the examples in the context embeddings. The input sequence embedding is represented as: 𝐸 = {𝐼_1_, …, *I_j_*, 𝑣_1_, …, 𝑣_*i*_, …, *e_n_*, …, *e_n_*} where 𝑣_*i*_ denotes the soft prompt vectors, which is randomly initialized from the embedding layer 𝑀 of LLM and is learnable during the training process. Here, the soft prompt vectors 𝑣_*i*_ serve as the traditional cue that aims to add additional example-relevant information, and bridge the task description with the examples, potentially enhancing the LLM’s ability to generalize from the provided examples to new instances.

The relation 𝑦 of entities pair in the input sentence is generated by the function 𝑝(𝑦|𝑥) = 𝑃(𝑦|𝑥, 𝐸) where 𝐸 is the embedding of demonstration, 𝑥 is the input sentence, and the text y is the desire output (relation types) for the input sentence x.

The objective of LEAP framework is 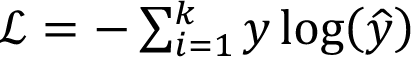 where *ŷ* represents the true label. 𝑦: represents the generated text of LEAP.

### Experiments and evaluation

During our research, we engaged with a suite of sophisticated LLMs. Noteworthy among these is Llama 2 [38], an innovative open-source language model from Meta AI acclaimed for its distinguished performance across numerous benchmarks, encompassing reasoning, coding, language mastery, and information retrieval tasks. We incorporated two models from the Llama 2 in our study: the Llama2-7b with 7 billion parameters, and the more expansive Llama2-13b with 13 billion parameters. We also employed MedLLaMA_13B [39], a specialized version of the Llama2-13b tailored with a Medical Corpus to enhance its medical query response capabilities.

Moreover, we included OpenAI’s GPT3.5 Turbo (hereafter referred to as GPT3.5) and GPT4 in our arsenal, both being LLMs celebrated for their remarkable language understanding and generation abilities.

In particular, we leveraged Llama2-7b [38], MedLLaMA_13B [39], GPT3.5, and GPT4 for the Demonstration Diversity Operation and selected Llama2-7b, Llama2-13b, and MedLLaMA_13B for Instruction-Example Adaptive Prompting. These models are at the forefront of language model architecture and are especially appropriate for the intricate task of relation extraction.

In evaluating the effectiveness of our LEAP framework, we benchmarked against a number of formidable baselines, including:

1. The BERT Family: A series of BERT variants, namely PubmedBERT [2], BioBERT [3], SciBERT [5], and BiolinkBert [6], were assessed for their performance on the datasets used in our research.
2. The Llama 2 Family: This includes the aforementioned Llama 2 models and the MedLLaMA_13B [39], which have demonstrated superior performance across various benchmarks. We applied instruction tuning to the *Instruction + Example* demonstration mode, without incorporating additional soft vectors. The approach is depicted in Figure 3a.
3. OpenAI’s GPT Models: The GPT3.5 and GPT4 served as baselines, noted for their comprehensive language processing capabilities.

For the tuning of LEAP, we experimented with the insertion of 5 to 20 soft tokens and tested learning rates of 1e-4 and 1e-5. Training was conducted with batch sizes ranging from 1 to 4. Our chosen evaluation metric was the micro-averaged F1 score, which provides a harmonic mean of precision and recall.

## RESULTS

The comparative analysis presented in Figure 4 delineates the performance differentials across various demonstration modes. The experiment’s performance metrics highlighted the superior efficacy of certain demonstration modes. Specifically, the *Instruction + Options + examples* and *Instruction + Options + options description+ examples* mode led to a notable increase in F1-scores across most models when compared to the baseline *Instruction + Options* mode. In the BC5CDR dataset, incorporating examples or options descriptions proved beneficial, with the mode *Instruction + options + options description + example* particularly standing out in the Llama2-13b model, achieving an impressive F1 score of 0.5070. Adding only examples was generally more advantageous than just adding the options description across all the models. For instance, the Llama2-7b model saw an F1 score increase to 0.353 from 0.073 with the addition of examples, while adding the *options description* increase to 0.215. Similarly, In the DDI 2013 dataset, the inclusion of examples consistently improved performance across the models, while the addition of options description had a more variable impact. Specifically, the *instruction + options + example* mode excelled in the Medllama*_*13B model, boosting the F1 score to an impressive 0.359. For larger models such as GPT3.5 and GPT4 in BC5CDR and DDI 2013, there was a noticeable trend where the more information provided, the better the models performed. The *instruction + options + example + options explanation* mode was best for GPT4, which achieved a peak F1 score of 0.770 in BC5CDR and 0.760 in DDI 2013, underscoring its ability to utilize detailed prompts effectively.

**Figure 4.**
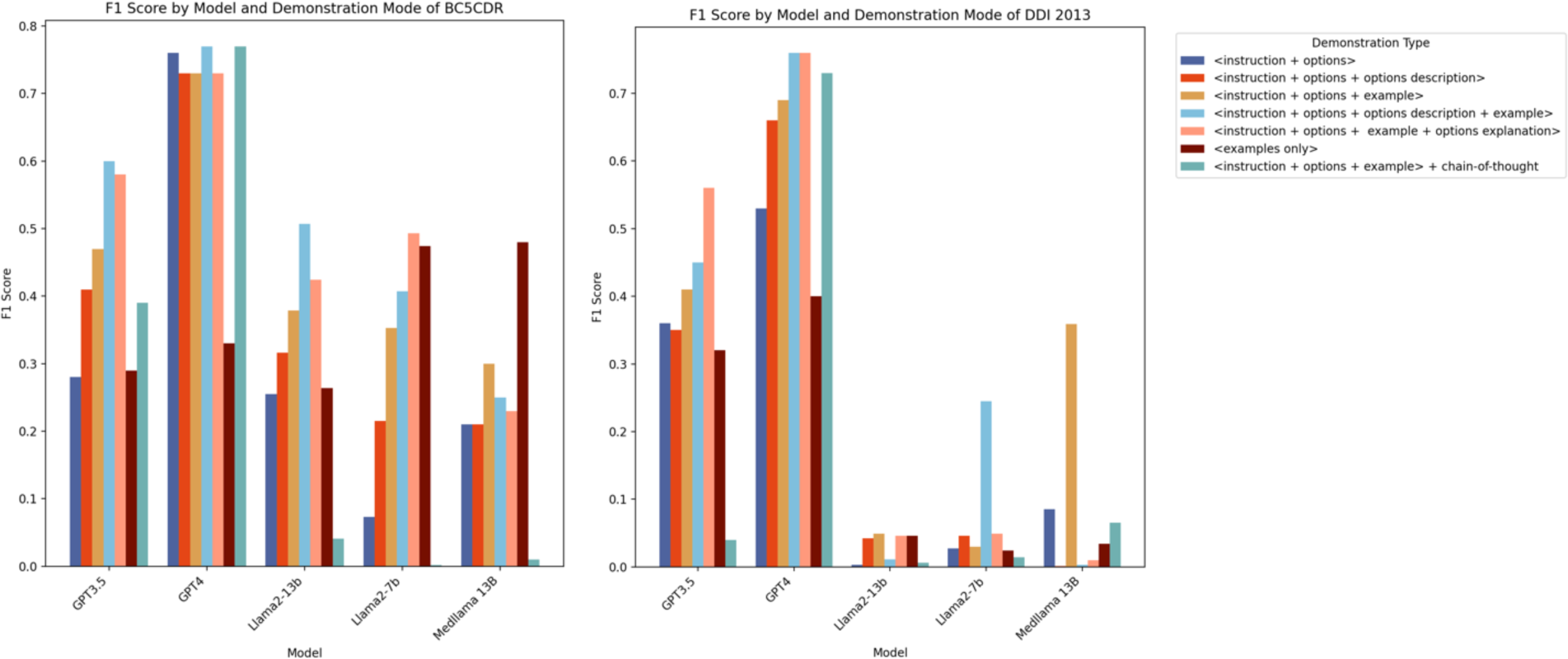
performance of demonstration diversity operation.

The *Chain-of-Thought prompt* mode exhibits varied effectiveness, with its impact differing significantly across models and datasets. For example, in the BC5CDR dataset, this demonstration mode did not lead to performance improvements for Llama2-7b and GPT3.5, where the F1 scores remained relatively low at 0.0020 and 0.3900, respectively. However, MedLLaMA_13B showed a substantial boost with a 50% increase in the F1 score when using the Chain-of-Thought mode, suggesting that this model may be better at processing and benefiting from the step-by-step reasoning this prompt structure provides. In the DDI 2013 dataset, the results were similarly mixed. The Chain-of-Thought prompts led to minimal changes for Llama2-7b and Llama2-13b, with F1 scores of 0.014 and 0.006, respectively, which does not represent a clear advantage over other prompt types. Yet, for MedLLaMA_13B, the Chain-of-Thought prompt resulted in an improvement, with an F1 score of 0.065, although this was not as high as the increase seen with the examples-only prompts.

Table 1 offers a detailed performance for two distinct tuning approaches: the instruction adaptive tuning and the example adaptive tuning. These strategies demonstrate that fine-tuning LLMs (Llama2-7b, Llama2-13b, MedLLaMA_13B) and employing adaptive tuning can significantly surpass the performance of traditionally fine-tuned LMs (BioBERT, SciBERT, BioLinkBERT, PubmedBERT) and even zero-shot GPT4, as evidenced by the metrics provided. Specifically, in the BC5CDR dataset, MedLLaMA_13B using example adaptive tuning reached the pinnacle of performance, boasting an F1 score of 95.13, while in the DDI 2013 dataset, Llama2-13b employing instruction adaptive tuning achieved the top F1 score of 92.24.

**Table 1.** Performance of RE in BC5CDR and DDI 2013.

| Method | Model | BC5CDR |  |  | DDI 2013 |  |  |
| --- | --- | --- | --- | --- | --- | --- | --- |
|  |  | Micro P | Micro R | Micro F1 | Micro P | Micro R | Micro F1 |
| Fine-tuning | BioBERT | 79.42 | 79.42 | 79.42 | 85.68 | 85.68 | 85.68 |
|  | SciBERT | 82.82 | 82.82 | 82.82 | 85.98 | 85.98 | 85.98 |
|  | BioLinkBERT | 83.54 | 83.54 | 83.54 | 85.88 | 85.88 | 85.88 |
|  | PubmedBERT | 82.36 | 82.36 | 82.36 | 85.31 | 85.31 | 85.31 |
| Zero-shot | GPT4 | 76.00 | 76.00 | 76.00 | 77.00 | 77.00 | 77.00 |
| Instruction tuning | Llama2-7b | 89.93 | 89.93 | 89.93 | 89.44 | 89.44 | 89.44 |
|  | Llama2-13b | 91.35 | 91.35 | 91.35 | 91.73 | 91.73 | 91.73 |
|  | MedLLaMA_13B | 92.70 | 92.70 | 92.70 | 91.33 | 91.33 | 91.33 |
| LEAP method 1 | Llama2-7b IAT* | 90.11 | 90.11 | 90.11 | 89.40 | 89.40 | 89.40 |
|  | Llama2-13b IAT | 85.53 | 85.53 | 85.53 | **92.24** | **92.24** | **92.24** |
|  | MedLLaMA_13B IAT | 91.44 | 91.44 | 91.44 | 91.91 | 91.91 | 91.91 |
| LEAP method 2 | Llama2-7b EAT* | 92.04 | 92.04 | 92.04 | 90.80 | 91.12 | 91.12 |
|  | Llama2-13b EAT | 94.70 | 94.70 | 94.70 | 91.76 | 91.76 | 91.76 |
|  | MedLLaMA_13B EAT | **95.13** | **95.13** | **95.13** | 92.20 | 92.20 | 92.20 |

\* IAT: instruction adaptive tuning, EAT: example adaptive tuning

In the BC5CDR dataset, the example adaptive tuning strategy, especially when applied to MedLLaMA_13B, significantly outperformed the fine-tuned models, achieving a superior F1 score of 95.13 compared to 92.70 for the fine-tuned version. Llama2-7b with its example adaptive tuning and Llama2-13b with its example adaptive tuning also reached notable high performance, with F1 score of 92.04 and 94.70, respectively, surpassing the results of instruction tuning, where Llama2-7b and Llama2-13b scored 89.93 and 91.35. Similarly, in the DDI 2013 dataset, the example adaptive tuning maintained superiority over the fine-tuning approach for all models tested. The example adaptive tuning also demonstrated a slight edge over the fine-tuning approach in models MedLLaMA_13B and Llama2-13b, affirming the potential of adaptive tuning in enhancing LLM performance.

While Instruction adaptive tuning exhibited fluctuating outcomes across both datasets. In the BC5CDR dataset, its advantage was observed solely in the Llama2-7b model, where it outperformed the same model utilizing standard instruction tuning. In the case of the DDI 2013 dataset, the deployment of instruction adaptive tuning on both Llama2-13b and MedLLaMA_13B models yielded better results than when these models were subjected to conventional instruction tuning.

## DISCUSSION

The discussion of the potential applications of generative LLMs in RE has gained momentum, as current research often gravitates towards deploying state-of-the-art methodologies CoT reasoning or retrieval-based techniques to construct hard instructions [21–29]. These methods aim to enhance generative LLMs’ ability to comprehend and process complex data. Notably, there is emerging evidence suggesting that prefix prompts may surpass the performance of fine-tuned models in clinical RE [40]. However, the literature still lacks a comprehensive exploration of adaptive tuning in demonstration and, more specifically, example adaptive tuning with the latest and most sophisticated generative LLM frameworks, such as Llama.

In the initial phase of our study, we have delved into various demonstration paradigms, seeking to understand their influence on RE tasks. With the *Instruction + Options + Examples* and *Instruction + Options + Examples +Options Description* modes leading to a notable increased performance, our findings resonate with the insights garnered from our prior research [27], indicating that the integration of examples within demonstration is not merely beneficial but critical for the success of RE tasks. This alignment underscores the significance of tailored examples in enhancing model comprehension and suggests a pivotal role for example-based learning in LLMs’ operational frameworks.

Moving into the second stage, our results show that the example adaptive tuning configuration yields superior performance for MedLLaMA_13B, Llama2-7b, and Llama2-13b models when juxtaposed with the instruction finetuning versions of the same models. This enhancement in performance indicates that example-based learning may provide a more robust framework for these models to understand and process complex tasks.

In the BC5CDR and DDI 2013 dataset, the instruction adaptive tuning’s performance varied among the models and did not yield the same level of enhancement as the example adaptive tuning. This variation suggests that within the domain of biomedical RE, prompts augmented with well-crafted examples are more critical than mere task explanations. This aligns with our initial phase findings, where the performance did not significantly improve with the addition of explanations for various relation labels compared to the mode adding the examples, reaffirming the importance of examples in RE task.

The implications of these findings are far-reaching. They invite us to reconsider the traditional reliance on extensive finetuning in favor of more dynamic, example-rich, and context-aware prompting strategies. By embracing this shift, we could potentially streamline the training process, reduce computational overheads, and accelerate the deployment of generative LLMs in real-world medical applications. Further research is warranted to validate these preliminary results and to explore the scalability of these approaches, potentially leading to a new paradigm in the training and application of generative LLMs for specialized tasks such as medical relation extraction.

This study, while offering valuable insights, does acknowledge certain constraints: Firstly, the scope did not extend to assessing the impact that varying lengths of soft prompts may have. Second, a comprehensive examination of how model size influences outcomes was not conducted. For instance, we did not implement the Llama 70b model in our experiments.

## CONCLUSION

Our investigation into the application of LLMs for RE tasks has unveiled promising avenues for enhancing model performance through LEAP adaptive tuning strategies. The study’s comparative analysis between instruction adaptive and example adaptive tuning on state-of-the-art models like Llama highlights the substantial benefits of incorporating contextually rich examples into demonstration. Our findings not only corroborate previous research advocating for example-centric approaches but also showcase the potential of these strategies to outperform traditional finetuning methods.

## CONTRIBUTORSHIP STATEMENT

HZ designed the study and drafted the manuscript. HZ, ML, and YX executed the experiments. All authors reviewed and finalized the manuscript.

## Data Availability

All data produced in the present work are contained in the manuscript

## ACKNOWLEDGEMENTS

We would also like to acknowledge to the staff at BPIC of University of Minnesota and the programmers from Rui Zhang’s group for their technical support.

## REFERENCES

1. Wang, Y., Wang, L., Rastegar-Mojarad, M., Moon, S., Shen, F., Afzal, N., Liu, S., Zeng, Y., Mehrabi, S., Sohn, S., & Liu, H. (2018). Clinical information extraction applications: A literature review. Journal of Biomedical Informatics, 77, 34–49. 10.1016/j.jbi.2017.11.011

2. Gu, Y., Tinn, R., Cheng, H., Lucas, M., Usuyama, N., Liu, X., Naumann, T., Gao, J., & Poon, H. (2022). Domain-Specific Language Model Pretraining for Biomedical Natural Language Processing. ACM Transactions on Computing for Healthcare, 3(1), 1–23. 10.1145/3458754

3. Lee, J., Yoon, W., Kim, S., Kim, D., Kim, S., So, C. H., & Kang, J. (2020). BioBERT: A pre-trained biomedical language representation model for biomedical text mining. Bioinformatics, 36(4), 1234–1240. 10.1093/bioinformatics/btz682

4. Roy, A., & Pan, S. (2021). Incorporating medical knowledge in BERT for clinical relation extraction. Proceedings of the 2021 Conference on Empirical Methods in Natural Language Processing, 5357–5366. 10.18653/v1/2021.emnlp-main.435

5. Beltagy, I., Lo, K., & Cohan, A. (2019). SciBERT: A Pretrained Language Model for Scientific Text. Proceedings of the 2019 Conference on Empirical Methods in Natural Language Processing and the 9th International Joint Conference on Natural Language Processing (EMNLP-IJCNLP), 3613–3618. 10.18653/v1/D19-1371

6. Yasunaga, M., Leskovec, J., & Liang, P. (2022). LinkBERT: Pretraining Language Models with Document Links. Proceedings of the 60th Annual Meeting of the Association for Computational Linguistics (Volume 1: Long Papers), 8003–8016. 10.18653/v1/2022.acl-long.551

7. Zhou, H., Austin, R., Lu, S.-C., Silverman, G. M., Zhou, Y., Kilicoglu, H., Xu, H., & Zhang, R. (2023). Complementary and Integrative Health Information in the literature: Its lexicon and named entity recognition. Journal of the American Medical Informatics Association, ocad216. 10.1093/jamia/ocad216

8. Clusmann, J., Kolbinger, F. R., Muti, H. S., Carrero, Z. I., Eckardt, J.-N., Laleh, N. G., Löffler, C. M. L., Schwarzkopf, S.-C., Unger, M., Veldhuizen, G. P., Wagner, S. J., & Kather, J. N. (2023). The future landscape of large language models in medicine. Communications Medicine, 3(1), 141. 10.1038/s43856-023-00370-1

9. Demszky, D., Yang, D., Yeager, D. S., Bryan, C. J., Clapper, M., Chandhok, S., Eichstaedt, J. C., Hecht, C., Jamieson, J., Johnson, M., Jones, M., Krettek-Cobb, D., Lai, L., JonesMitchell, N., Ong, D. C., Dweck, C. S., Gross, J. J., & Pennebaker, J. W. (2023). Using large language models in psychology. Nature Reviews Psychology, 2(11), 688– 701. 10.1038/s44159-023-00241-5

10. Li, M., Chen, M., Zhou, H., & Zhang, R. (2023). PeTailor: Improving Large Language Model by Tailored Chunk Scorer in Biomedical Triple Extraction. 10.48550/ARXIV.2310.18463

11. Mbakwe, A. B., Lourentzou, I., Celi, L. A., Mechanic, O. J., & Dagan, A. (2023). ChatGPT passing USMLE shines a spotlight on the flaws of medical education. PLOS Digital Health, 2(2), e0000205. 10.1371/journal.pdig.0000205

12. Singhal, K., Azizi, S., Tu, T., Mahdavi, S. S., Wei, J., Chung, H. W., Scales, N., Tanwani, A., Cole-Lewis, H., Pfohl, S., Payne, P., Seneviratne, M., Gamble, P., Kelly, C., Babiker, A., Schärli, N., Chowdhery, A., Mansfield, P., Demner-Fushman, D., … Natarajan, V. (2023). Large language models encode clinical knowledge. Nature, 620(7972), 172–180. 10.1038/s41586-023-06291-2

13. Tang, L., Sun, Z., Idnay, B., Nestor, J. G., Soroush, A., Elias, P. A., Xu, Z., Ding, Y., Durrett, G., Rousseau, J. F., Weng, C., & Peng, Y. (2023). Evaluating large language models on medical evidence summarization. Npj Digital Medicine, 6(1), 158. 10.1038/s41746-023-00896-7

14. Thirunavukarasu, A. J., Ting, D. S. J., Elangovan, K., Gutierrez, L., Tan, T. F., & Ting, D. S. W. (2023). Large language models in medicine. Nature Medicine, 29(8), 1930– 1940. 10.1038/s41591-023-02448-8

15. Zhang, S., Dong, L., Li, X., Zhang, S., Sun, X., Wang, S., Li, J., Hu, R., Zhang, T., Wu, F., & Wang, G. (2023). Instruction Tuning for Large Language Models: A Survey. 10.48550/ARXIV.2308.10792

16. Lou, R., Zhang, K., & Yin, W. (2023). Is Prompt All You Need? No. A Comprehensive and Broader View of Instruction Learning. 10.48550/ARXIV.2303.10475

17. Wei, J., Bosma, M., Zhao, V. Y., Guu, K., Yu, A. W., Lester, B., Du, N., Dai, A. M., & Le, Q. V. (2021). Finetuned Language Models Are Zero-Shot Learners. 10.48550/ARXIV.2109.01652

18. Li, M., & Huang, L. (2023). Understand the Dynamic World: An End-to-End Knowledge Informed Framework for Open Domain Entity State Tracking. 10.48550/ARXIV.2304.13854

19. Prasad, A., Hase, P., Zhou, X., & Bansal, M. (2023). GrIPS: Gradient-free, Edit-based Instruction Search for Prompting Large Language Models. Proceedings of the 17th Conference of the European Chapter of the Association for Computational Linguistics, 3845–3864. 10.18653/v1/2023.eacl-main.277

20. Zhou, Y., Muresanu, A. I., Han, Z., Paster, K., Pitis, S., Chan, H., & Ba, J. (2022). Large Language Models Are Human-Level Prompt Engineers. 10.48550/ARXIV.2211.01910

21. Wei, J., Wang, X., Schuurmans, D., Bosma, M., Ichter, B., Xia, F., Chi, E., Le, Q., & Zhou, D. (2022). Chain-of-Thought Prompting Elicits Reasoning in Large Language Models. 10.48550/ARXIV.2201.11903

22. Wan, Z., Cheng, F., Mao, Z., Liu, Q., Song, H., Li, J., & Kurohashi, S. (2023). GPT-RE: In-context Learning for Relation Extraction using Large Language Models. 10.48550/ARXIV.2305.02105

23. Chen, F., & Feng, Y. (2023). Chain-of-Thought Prompt Distillation for Multimodal Named Entity Recognition and Multimodal Relation Extraction. 10.48550/ARXIV.2306.14122

24. Wadhwa, S., Amir, S., & Wallace, B. C. (2023). Revisiting Relation Extraction in the era of Large Language Models. 10.48550/ARXIV.2305.05003

25. Meng, S., Hu, X., Liu, A., Li, S., Ma, F., Yang, Y., & Wen, L. (2023). RAPL: A Relation-Aware Prototype Learning Approach for Few-Shot Document-Level Relation Extraction (arXiv:2310.15743). arXiv. http://arxiv.org/abs/2310.15743

26. Xu, X., Zhu, Y., Wang, X., & Zhang, N. (2023). How to Unleash the Power of Large Language Models for Few-shot Relation Extraction? 10.48550/ARXIV.2305.01555

27. Li, M., Chen, M., Zhou, H., & Zhang, R. (2023). PeTailor: Improving Large Language Model by Tailored Chunk Scorer in Biomedical Triple Extraction. 10.48550/ARXIV.2310.18463

28. Gao, C., Fan, X., Sun, J., & Wang, X. (2023). PromptRE: Weakly-Supervised Document-Level Relation Extraction via Prompting-Based Data Programming. 10.48550/ARXIV.2310.09265

29. Rubin, O., Herzig, J., & Berant, J. (2021). Learning To Retrieve Prompts for In-Context Learning. 10.48550/ARXIV.2112.08633

30. Zhang, S., Dong, L., Li, X., Zhang, S., Sun, X., Wang, S., Li, J., Hu, R., Zhang, T., Wu, F., & Wang, G. (2023). Instruction Tuning for Large Language Models: A Survey. 10.48550/ARXIV.2308.10792

31. Liu, X., Ji, K., Fu, Y., Tam, W., Du, Z., Yang, Z., & Tang, J. (2022). P-Tuning: Prompt Tuning Can Be Comparable to Fine-tuning Across Scales and Tasks. Proceedings of the 60th Annual Meeting of the Association for Computational Linguistics (Volume 2: Short Papers), 61–68. 10.18653/v1/2022.acl-short.8

32. Lester, B., Al-Rfou, R., & Constant, N. (2021). The Power of Scale for Parameter-Efficient Prompt Tuning. 10.48550/ARXIV.2104.08691

33. Liu, X., Ji, K., Fu, Y., Tam, W. L., Du, Z., Yang, Z., & Tang, J. (2021). P-Tuning v2: Prompt Tuning Can Be Comparable to Fine-tuning Universally Across Scales and Tasks. 10.48550/ARXIV.2110.07602

34. Schick, T., & Schütze, H. (2021). Exploiting Cloze-Questions for Few-Shot Text Classification and Natural Language Inference. Proceedings of the 16th Conference of the European Chapter of the Association for Computational Linguistics: Main Volume, 255–269. 10.18653/v1/2021.eacl-main.20

35. Brown, T. B., Mann, B., Ryder, N., Subbiah, M., Kaplan, J., Dhariwal, P., Neelakantan, A., Shyam, P., Sastry, G., Askell, A., Agarwal, S., Herbert-Voss, A., Krueger, G., Henighan, T., Child, R., Ramesh, A., Ziegler, D. M., Wu, J., Winter, C., … Amodei, D. (2020). Language Models are Few-Shot Learners. 10.48550/ARXIV.2005.14165

36. Herrero-Zazo, M., Segura-Bedmar, I., Martínez, P., & Declerck, T. (2013). The DDI corpus: An annotated corpus with pharmacological substances and drug–drug interactions. Journal of Biomedical Informatics, 46(5), 914–920. 10.1016/j.jbi.2013.07.011

37. Taboureau, O., Nielsen, S. K., Audouze, K., Weinhold, N., Edsgard, D., Roque, F. S., Kouskoumvekaki, I., Bora, A., Curpan, R., Jensen, T. S., Brunak, S., & Oprea, T. I. (2011). ChemProt: A disease chemical biology database. Nucleic Acids Research, 39(Database), D367–D372. 10.1093/nar/gkq906

38. Touvron, H., Martin, L., Stone, K., Albert, P., Almahairi, A., Babaei, Y., Bashlykov, N., Batra, S., Bhargava, P., Bhosale, S., Bikel, D., Blecher, L., Ferrer, C. C., Chen, M., Cucurull, G., Esiobu, D., Fernandes, J., Fu, J., Fu, W., … Scialom, T. (2023). Llama 2: Open Foundation and Fine-Tuned Chat Models. 10.48550/ARXIV.2307.09288

39. Chaoyi-wu/MedLLaMA_13B · Hugging Face. (n.d.). Retrieved December 13, 2023, from https://huggingface.co/chaoyi-wu/MedLLaMA_13B

40. Peng, C., Yang, X., Smith, K. E., Yu, Z., Chen, A., Bian, J., & Wu, Y. (2023). Model Tuning or Prompt Tuning? A Study of Large Language Models for Clinical Concept and Relation Extraction. 10.48550/ARXIV.2310.0623

